# Navigating stroke care during the COVID-19 pandemic: A scoping review

**DOI:** 10.1101/2024.03.19.24304563

**Authors:** Alixe Ménard, Michelle Yang, Samantha Oostlander, Kanika Sarwal, Irfan Manji, Tracey O’Sullivan, Sarah Fraser

## Abstract

**Background:** Previous viral outbreaks have highlighted implications for the management of complex health conditions. This study delves into the repercussions of the COVID-19 pandemic on stroke care, by examining evidence of shifts in healthcare utilization, the enduring effects on post-stroke recovery, and the overall quality of life experienced by stroke survivors.

**Methods:** A scoping review was conducted following the Joanna Briggs Institute Methodology for Scoping Reviews. The search strategy encompassed electronic databases (APA PsycInfo, Embase, Medline, and CINAHL). English language articles published between December 2019 and January 2022 were included, focusing on individuals who experienced a stroke during the COVID-19 pandemic. Data extraction involved identifying study characteristics and significant findings, facilitating a qualitative and narrative synthesis of the gathered evidence.

**Results:** Seven domain summaries were identified. They all described the aspects of systemic transformations in stroke care during the COVID-19 pandemic: (1) patient behavior and awareness; (2) telemedicine and remote care; (3) delays in treatment; (4) impact on healthcare resources; (5) quality of care; (6) changes in stroke severity; and (7) reduction in stroke admissions.

**Conclusions:** This study underscored the critical need to encourage swift patient response to acute stroke symptoms, by finding new avenues for treatment, mitigating hospital-related infection fears, and advocating for the establishment of centralized stroke centers. These measures are integral to optimizing stroke care delivery and ensuring timely interventions, particularly in the challenging context of a pandemic.

## INTRODUCTION

Past large-scale outbreaks, exemplified by the 2015 Middle East Respiratory Syndrome (MERS), have been associated with changes in the utilization of emergency care services for conditions such as ischemic stroke and myocardial infarctions^1^. Similarly, the COVID-19 outbreak, as a result of SARS-CoV-2, demonstrated an impact on the management of cardiovascular disease^2^. The World Health Organization (WHO) emphasizes the necessity of drug treatment for hypertension, diabetes, and hyperlipidemia in effectively managing cardiovascular diseases^3^. Successful cardiovascular management requires patients to be involved in their care, having social and interprofessional support, and consistent follow-ups with healthcare providers^4^. Although these comprehensive approaches to stroke are crucial, when COVID-19 spread across regions worldwide, there were escalating concerns regarding the capacity of global healthcare systems to meet rising demands for acute emergencies and provide effective rehabilitation services for individuals who experienced strokes^5^. In this review, we assess the global impact of the COVID-19 disaster on stroke care, focusing on evidence for changes in healthcare access and utilization, long-term impacts on post-stroke recovery, and the quality of life of stroke survivors.

The WHO declared the COVID-19 pandemic on March 11^th^, 2020, catapulting many countries into a state of emergency^6^. Many institutions reported that this affected hospital resource allocation and well-established pathways for care^7^. This includes the protocols and process measures for the care/management of stroke^7^, the second leading cause of death and the foremost cause of disability worldwide^8,9^.

Each year, 15 million individuals around the globe experience a stroke^10^. To address the challenges posed by COVID-19 on healthcare systems, novel strategies were introduced to enhance in-hospital workflow, optimize the availability of healthcare workers, and minimize risks for patients, families, and care partners^8,11^. These changes largely affected people experiencing stroke and other non-COVID-19 conditions, which required extensive resources related to imaging, diagnostics, treatment options, and length of stay^12^. The impact of the ongoing crisis on stroke care continues to be influenced by in-hospital decisions, regional public health protocols, prehospital triage, and equitable access to acute treatments^13^. It is also the result of different state-wide strategies to prevent and contain the spread of COVID-19^14^. These strategies (e.g. quarantine, isolation, screening and triage protocols), may not only influence traditional stroke pathways but also health-seeking behaviors^15^.

The negative impacts of preventative public health initiatives that target the spread of infections within hospitals have been recognized, and they are anticipated to give rise to various epidemiological consequences (e.g., increased mental health disorders, antibiotic resistance)^16^. As the pandemic persists, there will likely be a continued shift in the epidemiology of stroke and other cardiovascular events^17^. In Italy, the restrictive lockdown required adaptations of the integrated care pathways (ICP) serving stroke and other time-dependent conditions^18^. In the United Kingdom, non-urgent elective care was paused as the English National Health Service (NHS) reprioritized the delivery of care for COVID-19 patients^19^. In the United States, emergency treatment protocols, such as for thrombolysis and endovascular revascularization, were at risk of significant time delays^20^. The urgency imposed by COVID-19 pushed these rapid public health reforms. With minimal forewarning and planning, healthcare institutions were forced to swiftly formulate, communicate, and implement an organizational response to the pandemic. This involved recalibrating healthcare delivery to strike a balance between providing timely and effective care while mitigating the risk of COVID-19 infection. For example, the American Heart Association and American Stroke Association quickly issued temporary emergency guidelines for stroke hyperacute management^21^, and the Canadian Stroke Best Practice recommendations were rapidly modified to provide a virtual tele-stroke healthcare management-toolkit^22^.

During disasters, stroke survivors are likely to experience serious disruptions that can impact social, cognitive, and motor outcomes^23^. The impacts of the pandemic on stroke survivors produced “collateral damage” ^24^, yet the global evidence for the direct and indirect impacts of the COVID-19 pandemic on stroke care has yet to be synthesized. Hence, to support knowledge mobilization, the purpose of this scoping review is to form an evidence-based framework to describe the resiliency and capacity of stroke care systems during the pandemic and to formulate recommendations for reducing the risks for stroke patients during an emergency/disaster. This knowledge will inform future emergency preparedness protocols supporting effective response and highlight the needs of stroke survivors in a disaster context.

## METHODS

Scoping review methodology allows researchers to navigate the landscape of evidence by exploring emerging findings in situations where clarity is lacking, allowing for the formulation of more comprehensive and specific questions^25^. The value of conducting this study as a scoping review is that it will inform evidence-based practices in the broader field of cardiovascular healthcare service delivery by identifying gaps in the research knowledge base^25^. The Joanna Briggs Institute (JBI) Methodology for Scoping Reviews^26^, provides a contemporary methodological framework to facilitate knowledge translation in health research. This framework instructed our reporting outline, inclusion criteria, search strategy, extraction, presentation and summarization of the results, and any potential implications for research and practice.

### Search strategy

The following four electronic databases were systematically searched as shown in the PRISMA chart in Appendix A: APA PsycInfo, Embase, Medline, and CINAHL. Additionally, grey literature was hand-searched using open-access repositories and the reference lists from selected articles. Two levels of search terms were used: (1) terminology for stroke and (2) the COVID-19 pandemic. The protocol for the review design and search strategy was developed by study researchers (MY, SO) and health science librarians from the University of Ottawa Library (KF & NL) prior to database searches. The search strategy is presented in Appendix B. Guidelines from the Preferred Reporting Items for Systematic Reviews and Meta-Analyses extension for Scoping Reviews (PRISMA-ScR) were followed, and the PRISMA-ScR checklist, which guides the reporting of scoping reviews was completed^27^. This reporting guideline is consistent with the JBI guidance for scoping reviews and highlights the importance of methodological rigor in conducting scoping reviews^25^.

### Study screening and selection

All stages of the screening process (i.e., title and abstract screening, full text screening and selection) were completed using Covidence. Two researchers (MY, SO) participated independently at each stage. In the case of any discordance between authors’ selection, there was a discussion between co-authors (MY, SO, IM, AM & KS) to reach a consensus regarding the inclusion or exclusion of the article.

English language articles published between December 2019 (the inception of the COVID-19 pandemic) and January 2022 were included. Population characteristics include people who had a stroke (clinically defined as hemorrhagic stroke (intracerebral and subarachnoid), ischemic stroke, TIA, or any condition that can be classified as a cerebrovascular accident) during the COVID-19 pandemic, as defined by the period between when SARS-CoV-2 was initially identified as a threat - to January 2022. The outcomes of interest of selected articles were the impact of the COVID-19 pandemic on individuals who had a stroke, including stroke care during the pandemic, changes in healthcare delivery, functional outcomes, and quality of life after stroke. Articles presenting original research findings or analyses thereof were included, including primary and secondary data, or synthesis of available literature reflecting the functional outcomes of patients who had a stroke.

Studies were excluded if the outcomes of interest were the impact of SARS-CoV-2 on stroke symptoms, stroke as a complication or prognostic factor of SARS-CoV-2, or risk of stroke in people who contracted COVID-19. Opinion pieces, commentary papers and recommendation papers were excluded if they did not report on empirical results from an original study. Articles from the position of, or solely focusing on the experiences of, Health Care Professionals (HCPs) were excluded.

### Data collection and analysis

Authors (MY, IM, AM & KS) jointly developed a data-charting table to determine the variables to extract. Extraction was initially conducted by one author (MY) and confirmed for accuracy by other authors (IM, AM & KS). All authors iteratively updated the extraction tables. We extracted data on study characteristics (Appendix C), including year, country, study design, and significant findings from the manuscripts. A comprehensive qualitative and narrative synthesis of the gathered data was performed in preparation for a content analysis^28^. The authors refined the analysis and presentation through thoughtful deliberations and consensus-building, particularly when adjustments were required or in the presence of conflicting opinions. To establish coding reliability, title and abstracts were dually coded, followed by the full inclusion of articles. Title and abstract screening for interrater reliability was initially performed by MY and SO, resulting in a strong agreement with a kappa value of 0.85. Following this, a secondary screening of the 84 included articles was carried out by AM and KS, yielding a kappa value of 0.9, indicating strong agreement.

## RESULTS

### Study characteristics

The search and screening processes are presented in Appendix A. We identified a total of 861 studies after the removal of duplicate studies. A total of 650 records were excluded following title and abstract screening. Of the 211 full articles assessed for eligibility, 84 met inclusion criteria and were retained.

The included articles underwent a reevaluation by three members of the research team (KS, IM & AM) to further confirm their eligibility for inclusion. The included articles were composed of 46 retrospective studies (cohort (n=7); observational (n=11); cross-sectional (n=3); archive (n=1); review (n=1); unspecified (n=23)); 10 prospective studies (cohort (n=2); observational (n=3); cross-sectional (n=3); unspecified (n=2)); 26 observational studies (reports (n=9); cohort (n=7); cross-sectional (n=1); unspecified (n=9)); one matched pairs study; and one survey and one exploratory design. The included articles demonstrated wide geographical spread spanning Bangladesh^29^; Canada^30–32;^ Chile^33^; China^15,34–37,37–44;^ Egypt^45^; Finland^46^; France^47^; Germany^48–50;^ India^51^; Iran^52,53^; Israel^54–56;^ Italy^8,57–67;^ Japan^9,68^; Lithuania^69^; Malaysia^70^; Netherlands^71^; Norway^72,73^; Saudi Arabia^74,75^; Spain^76–79;^ Switzerland^80^; Taiwan^81^; United Kingdom^82–84;^ and United States of America^7,20,24,39,85–104^. The studies encompassed participants from a wide range of diverse backgrounds, including individuals from various racial and ethnic communities, linguistic minorities, and persons with differing levels of disability. A wide range of ages was also captured in the articles ranging from early to late adulthood.

### Review findings

A total of seven domain summaries were identified through content analysis^28^. This analytical approach embraces the consideration of multiple perspectives to gain a comprehensive understanding of a specific area of focus in a given timeframe (i.e., stroke care during the COVID-19 pandemic), through systematic content analysis^105^. After becoming familiar with the data, domains were developed, and summaries for each domain were compiled. The domain summaries reflect the diverse ways in which the COVID-19 pandemic affected stroke care globally. They highlight the challenges faced by healthcare systems in maintaining quality stroke care amid the disruptions caused by the pandemic. These domain summaries include: (1) patient behavior and awareness; (2) telemedicine and remote care; (3) delays in treatment; (4) impact on healthcare resources; (5) quality of care; (6) changes in stroke severity; and (7) reduction in stroke admissions. The domain summaries were then grouped into three domains: patient conduct, healthcare delivery, and stroke characteristics, as depicted in Figure 1.

**Figure 1.**
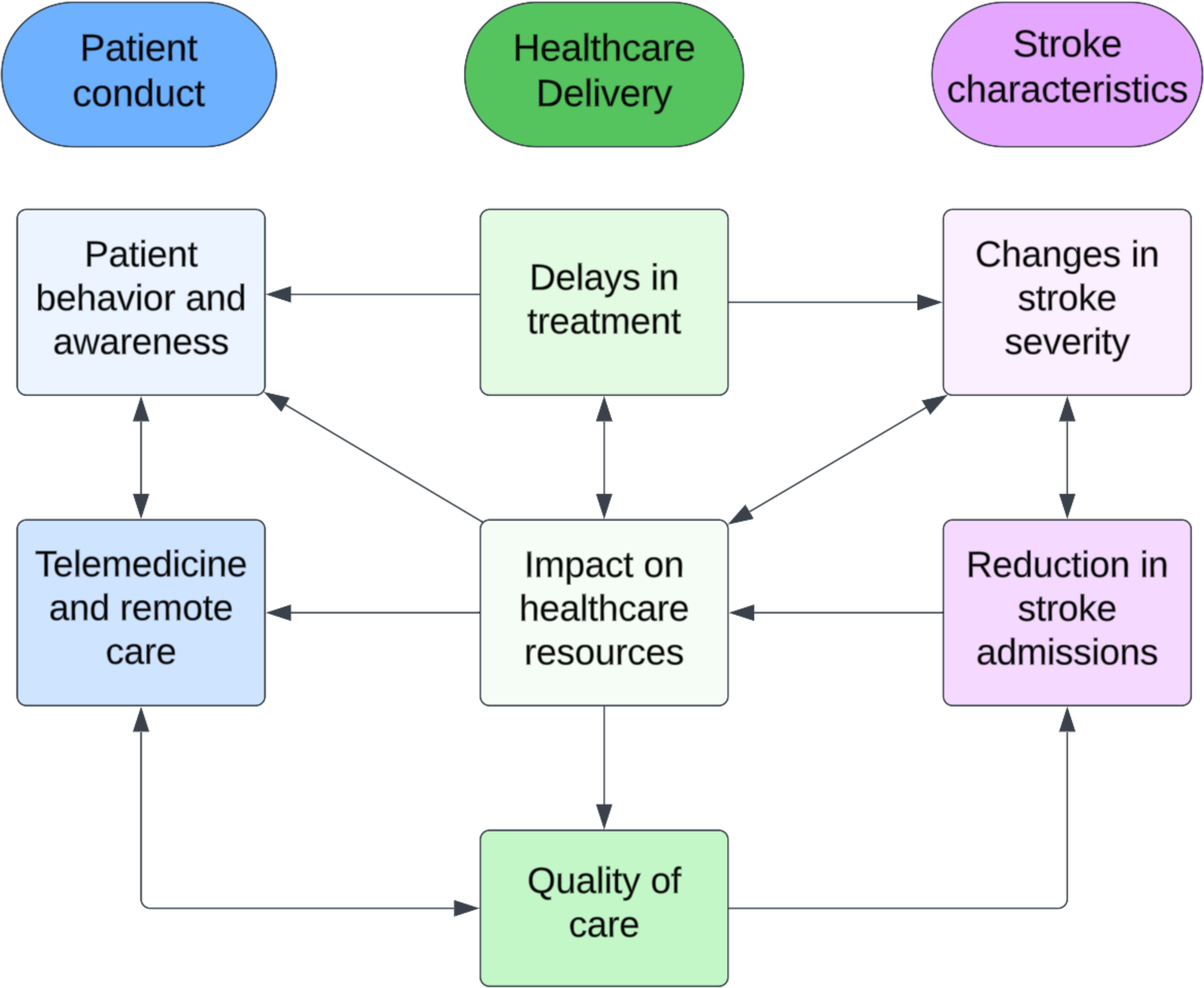
Systemic and interrelated transformations in stroke care during the COVID-19 pandemic presented as a framework.

### Domain 1: Patient conduct

#### Domain Summary 1: Patient behavior and awareness

Patient behavior, such as the reluctance to seek medical attention, emerged as a significant factor affecting stroke outcomes during the pandemic. The fear of contracting COVID-19 and changes in healthcare-seeking behavior were recurrent themes^42,77,96^. Individuals experiencing mild stroke symptoms and those with lower functional abilities were less inclined to seek medical attention^7,42,59,77^. Patients tended to delay seeking medical help, often surpassing the eligible treatment timeframes^33,39^. Additionally, external factors such as national traffic control measures (for example, in China) played a role in hindering prompt access to healthcare services^15^. Moreover, the implementation of additional COVID-19 prevention protocols, such as mandatory travel history checks, contributed to prolonged stroke management periods^15^.

#### Domain Summary 2: Telemedicine and remote care

Amid the pandemic, there was a surge in patient reliance on telemedicine for stroke consultations and follow-ups, particularly for non-acute treatment, as noted in numerous studies^31,77,79,87,90,92^. This shift in healthcare delivery aimed to address pandemic-related challenges and ensure ongoing patient care. However, despite these efforts, older adults experienced a reduction in weekly telestroke consultations and decreased utilization of telestroke services^24^. Conversely, there was a significant decline in in-person outpatient visits and inpatient care for stroke patients, particularly affecting high-risk populations such as immunocompromised individuals and those with underlying health conditions^31,77,79,87,90,92,99^. While telemedicine became increasingly common for both new and existing patients in younger cohorts, outpatient clinics that remained operational implemented stringent safety measures such as the use of personal protective equipment (PPE) and isolation protocols^36,37,42,46,87^.

### Domain 2: Healthcare delivery

#### Domain Summary 3: Delays in treatment

The focus on COVID-19 patients and facilities led to a restructuring of pre-hospital transportation, decreased interactions with stroke specialists, centralization of stroke centers and rehabilitation routes, a reduction in the availability of additional diagnostic tests, timely therapies, and emergency pathways for stroke patients^45,51,82^. These changes resulted in delays, which in turn affected the utilization of immediate treatments, and had implications for prognosis^76^. The advent of the COVID-19 pandemic prompted a shift in healthcare priorities, diverting attention and resources away from routine stroke care, including the essential procedures of thrombolysis and thrombectomy^20,45,72,104^. As a result, the pandemic caused a strain on healthcare systems, resulting in a noticeable decline in the performance of these two critical interventions by skilled practitioners such as cardiologists, radiologists, vascular surgeons and emergency medicine physicians^20,45,72,104^. Fewer referrals for specialized stroke care were made during the pandemic^48,55,69,75,83,98^. Factors contributing to these delays included changes in hospital protocols, reorganization of healthcare resources, and changes in patient health-seeking behaviors as a result of fear^42,77,96^. The crowding of hospitals during the COVID-19 pandemic not only led to delayed acute treatment for stroke patients,^75^ but also resulted in increased symptoms-to-door time (i.e., the time between the onset of symptoms and arrival at the hospital) and door-to-treatment time (i.e., the time between arrival at the hospital and the initiation of treatment) for reperfusion.^30,34,60,76,79^. However, prehospital triage proved helpful in the provision of timely care in resource-limited areas^69^. Notably, some hospital Emergency Departments implemented two distinct intra-hospital pathways to ensure the safe management of stroke patients^65^.

#### Domain Summary 4: Impact on healthcare resources

The reorganization of healthcare resources, reallocation of stroke professionals, and the prioritization of COVID-19 patients^59^ led to changes in stroke care pathways, delays in acute treatment^75^, and consequences for stroke outcomes^76^. Ambulance services experienced an excessive burden due to the pandemic^58^. There was also an increase in the demand for computed tomography (CT) scans^100^; discontinuation of rehabilitation programs due to the reorganization of resources; shortfalls in health insurance coverage for stroke patients; and limited access to specialized care units and diminished workforce in hospitals^59,68,74,102^. Ultimately, these pandemic-related healthcare interruptions led to a fragmented stroke chain of survival and care pathways.

#### Domain Summary 5: Quality of care

During the COVID-19 pandemic, reductions in quality-of-care metrics, including door-to-CT (time from arrival to hospital and initiation of a CT scan), dysphagia screenings, early antithrombotic use, and rehabilitation evaluations, were observed in various studies^70,77,81,83,90,99,100,102^. Despite discussions about the preservation and, in some cases, improvement in stroke care quality during the pandemic^20,81,82^, challenges and disruptions in outpatient care, rehabilitation, and follow-up care were noted in several articles^6,7,30,62,65,66^. Furthermore, studies indicated that patients treated during the pandemic had worse discharge outcomes and increased mortality rates^20,33,85^. Regrettably, higher rates of in-hospital death and hospice discharges were observed during the pandemic period, attributed to longer treatment delays^16,26,79,87^. Additionally, higher post-stroke disability (e.g., loss of peripheral vision, muscle spasticity, hemiplegia^106^), measured using the modified Rankin Scale, was correlated with the discontinuation of rehabilitation services due to the pandemic^74,102^.

### Domain 3: Stroke characteristics

#### Domain Summary 6: Changes in stroke severity

During the pandemic, noticeable shifts in stroke severity were observed, with many studies highlighting an escalation in the severity of strokes compared to pre-pandemic measures^33,40,48,63,91,98,103^. Particularly noteworthy is the finding that stroke severity upon hospital admission was significantly higher in the pandemic period compared to pre-pandemic cohorts^48,91^. This phenomenon may be attributable to delayed patient presentations; a lack of close observers to notice stroke symptoms due to lockdowns and stay-at-home directives; and a hesitancy among individuals to promptly seek medical attention.

#### Domain Summary 7: Reduction in stroke admissions

Many articles discussed the impact of the COVID-19 pandemic on hospital admissions for stroke patients^9,15,29,36,40,44,45,47–54,58,60,61,65,66,69,71,72,82,83,86,88,91,103,104^. A few articles mentioned a decrease in the number of stroke admissions during the pandemic^29,39,40,52,53,69,72,76,77,79,83,84,90,97,107^ despite reperfusion therapy, thrombolysis and thrombectomy rates remaining unchanged during this period^50,54,63,65,71,91^. There were also fewer presentations of emergent large vessel occlusion (ELVO) in the emergency departments^45^, less stroke unit admissions^8,64,79^, and fewer code strokes^77,79^. A reduction in transfers between facilities was also observed, likely attributed to precautionary measures implemented early in the pandemic to minimize the risk of COVID-19 transmission through interfacility transfers^57,98,108^. These reductions in stroke admissions were attributed to factors such as patient reluctance to seek medical care, fear of infection, and changes in healthcare protocols^61,77,82,96^.

## DISCUSSION

In this scoping review we aimed to explore the repercussions of the COVID-19 pandemic on stroke care, examining evidence concerning shifts in healthcare utilization, the enduring effects on post-stroke recovery, and the overall quality of life experienced by stroke survivors. A thorough literature search uncovered 84 articles that satisfied the inclusion criteria for this review. Included articles reflected the condition of stroke care at healthcare facilities or the stroke experiences of individuals from several continents. Findings from the articles were analyzed using content analysis^105^. This resulted in three domains: patient conduct, healthcare delivery and stroke characteristics; and seven domain summaries: (1) patient behavior and awareness; (2) telemedicine and remote care; (3) delays in treatment; (4) impact on healthcare resources; (5) quality of care; (6) changes in stroke severity; and (7) reduction in stroke admissions, as presented in Figure 1.

The results of the current review align with existing literature wherein pandemic-related health measures and states of emergency affect the likelihood of seeking medical care during acute cardiovascular incidents^109^. Current research suggests that states of emergency influence patient behavior by decreasing admission rates for cardiovascular events and causing broader shifts in health-seeking behaviors^109^. Similarly, results from this review indicate that patients feared contracting SARS-CoV-2, thus when making health-related decisions, the risk of being admitted to a hospital for an acute cardiovascular event was weighed against the risk of COVID-19 mortality for older and/or institutionalized patients^110^.

In sum, the impact of COVID-19 on stroke care was multifaceted, with social isolation and stay-at-home orders contributing to fewer witnesses of stroke symptoms, delayed recognition of mild stroke signs, and increased avoidance of advanced care^7,42,59,77^. Raising awareness about stroke symptoms and the critical need for immediate medical attention, even amidst a pandemic, is paramount^111^. This effort can encompass diverse strategies, including public service announcements, targeted social media Face, Arm, Speech, Time (FAST) campaigns during lockdowns, and proactive community outreach initiatives^112^. Additionally, fostering support networks through structured systems of assistance within communities to aid individuals experiencing stroke symptoms, particularly those who are socially isolated or face barriers in seeking assistance, is essential for mitigating adverse stroke outcomes during times of crisis^113^.

Amidst crises, stroke care systems face numerous challenges, including delays in hospital presentation, ineligibility for time-sensitive treatment, and a reduction in thrombolysis and thrombectomies^20,45,60,86^. To address these challenges, adaptability in stroke care systems is essential. Recent research has indicated that the successful implementation of performance improvement programs and robust stroke care systems can help maintain high-quality care during health emergencies^114^. Additionally, mitigating delays and preventing reluctance to seek medical care requires addressing various factors such as overcoming fear of COVID-19 infection^115^; managing transportation restrictions (i.e., many individuals indicated being ensure whether travel to hospital was permitted and feared exposing patients to COVID-19^116^), and alleviating burdens on the healthcare system. Notably, innovations have been proposed to alleviate these burdens and safeguard stroke survivors during crises, including: (1) implementing teleneurology networks to enhance thrombolysis access for acute ischemic stroke; (2) enhancing the efficiency of emergency care for acute ischemic stroke (e.g., by preregistering patients during transport via ambulance and having the neurologist obtain clinical history electronically prior to arrival at the hospital); and (3) offering alternatives to inpatient care for transient ischemic attacks through an accelerated diagnostic protocol in the observation unit^117^.

This emphasizes the importance of rapid adaptation and multidisciplinary changes in stroke care protocols, including effective triage screening, proper PPE utilization, and increased equipment cleaning to minimize treatment delays^118^. Recognizing the impact of social isolation and the stress of the pandemic on the mental health of patients and staff also underscores the need for comprehensive psychological support measures in stroke care strategies^119^. To address COVID-19-related fears, stroke care must include the integration of psychological support, particularly emphasizing the consideration of physiological risk factors linked to COVID-19 comorbidities^115^. For future preparedness, a holistic approach that encompasses technological advancements, public education, and mental health support will be essential to ensure optimal patient outcomes and maintain the integrity of stroke care systems during crises. Findings from this review also point to a need to compare the quality of stroke care delivered through telemedicine with that provided in traditional, in-person settings.

### Strengths and limitations

While the scoping review offers valuable insights into the challenges faced by stroke care systems and survivors during the COVID-19 pandemic, it is important to acknowledge certain limitations. One notable limitation is the restriction of the search to English language articles, which may have resulted in the exclusion of relevant studies published in other languages, potentially limiting the comprehensiveness of the review. However, by analyzing studies from multiple continents and considering diverse populations, the review provides a global perspective on the challenges faced by stroke care systems during the pandemic. Additionally, the review includes articles published only up to January 2022, which may not capture the most recent developments in stroke care during the ongoing pandemic. Furthermore, while the review offers a qualitative synthesis of the literature, the absence of quantitative analysis limits the ability to quantify the magnitude of the impact of COVID-19 on stroke care outcomes. Despite these limitations, the review provides a thorough examination of the impact of the COVID-19 pandemic on stroke care, encompassing various aspects such as changes in healthcare utilization, long-term effects on post-stroke recovery, and quality of life considerations for stroke survivors. The use of scoping review methodology, guided by the JBI framework, ensures a systematic and transparent approach to navigating the available evidence.

### Conclusions

As evidenced by our scoping review, the COVID-19 pandemic has profoundly altered stroke care around the globe. Heightened patient fear and subsequent delays in seeking medical attention, coupled with rapid shifts towards telemedicine, highlight the urgent need for adaptable stroke care systems. Recommendations include prioritizing telemedicine and improving its access to older adults and high-risk populations; and implementing robust performance improvement programs to mitigate treatment delays. Future research should explore strategies for enhancing patient education and communication to address fear-related delays in seeking medical attention. Longitudinal studies tracking the long-term effects of pandemic-induced changes in stroke care delivery are also needed for understanding their sustained impact and informing future emergency preparedness protocols.

## Data Availability

The data that support the findings of this study are available on request from the corresponding author, [AM].

## Acknowledgements

We thank librarians, Karine Fournier and Nigèle Langlois, from the University of Ottawa Health Sciences Library for their invaluable assistance with our comprehensive systematic search procedure.

## Appendix A. PRISMA chart of search and screening process

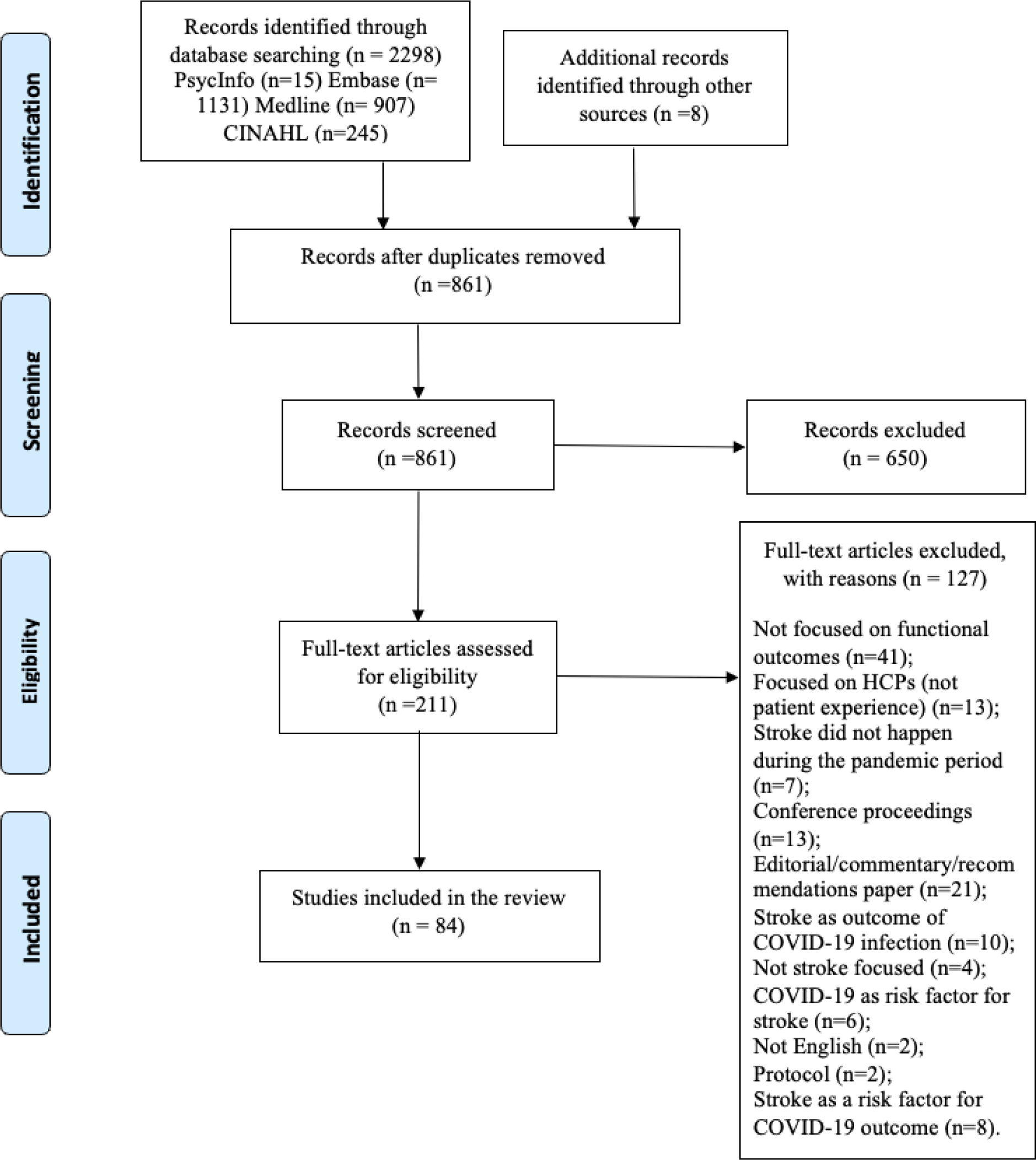

## Appendix B. Medline Search

1. exp Coronavirus/

2. exp Coronavirus Infections/

3. coronavirus*

4. corona virus*

5. OC43

6. NL63

7. 229E

8. HKU1

9. HCoV*

10. ncov*

11. covid*

12. sars-cov*

13. sarscov*

14. Sars-coronavirus*.mp

15. hemorrhagic stroke (intracerebral and subarachnoid)

16. ischemic stroke

17. Transient Ischemic Attack (TIA)

18. any condition that can be classified as a cerebrovascular accident

## Appendix C. Included studies exploring stroke care during the COVID-19 pandemic

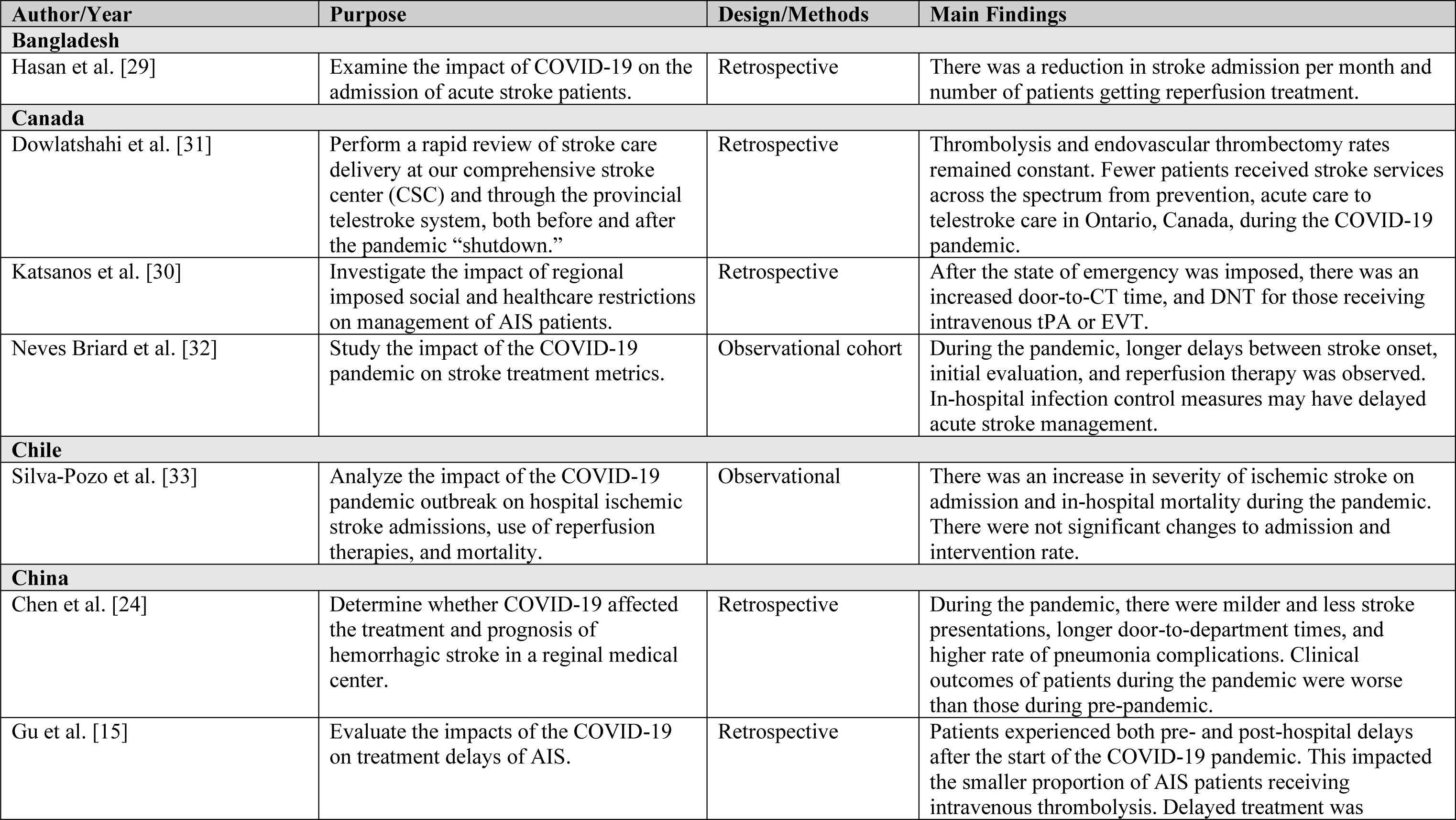
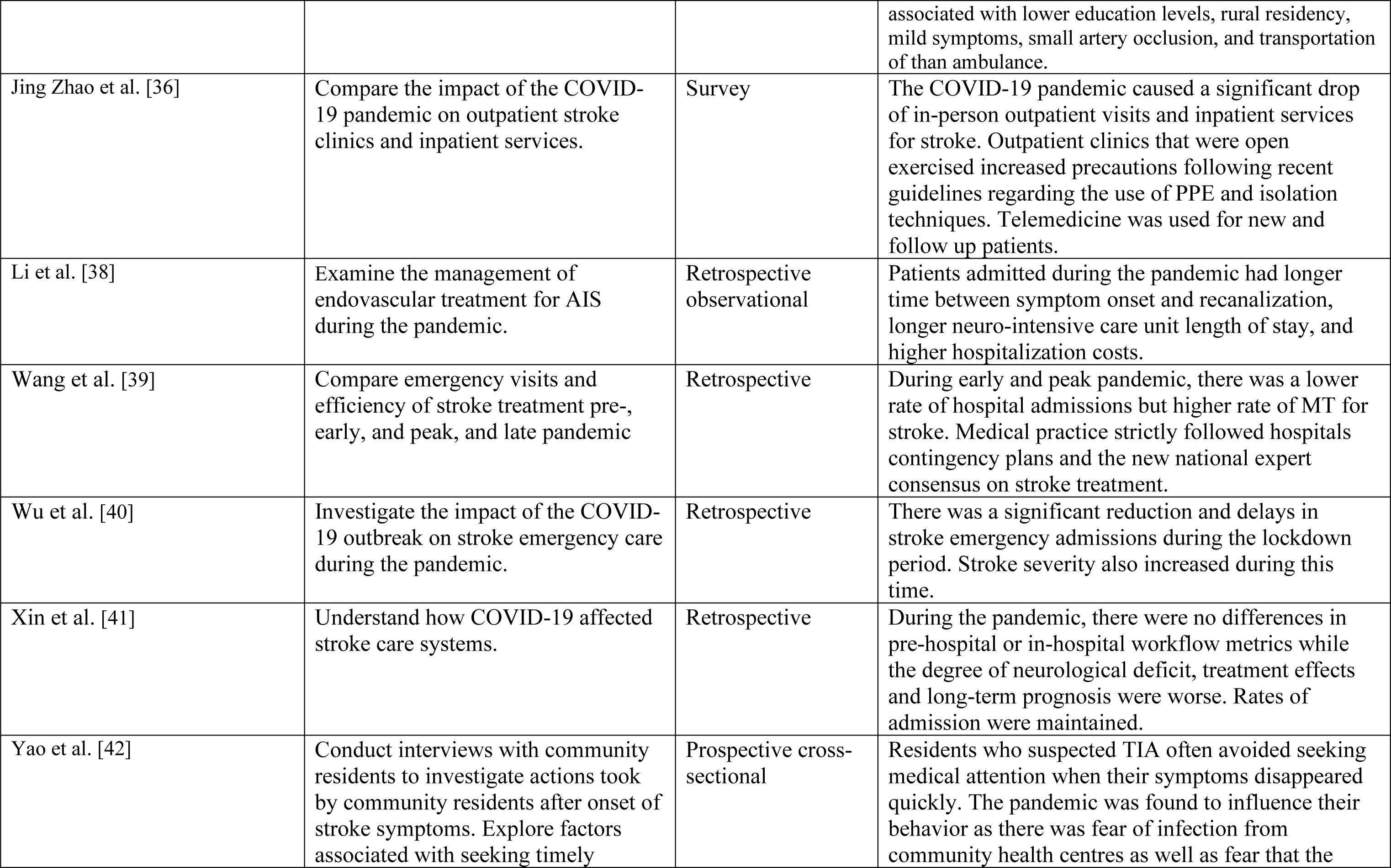
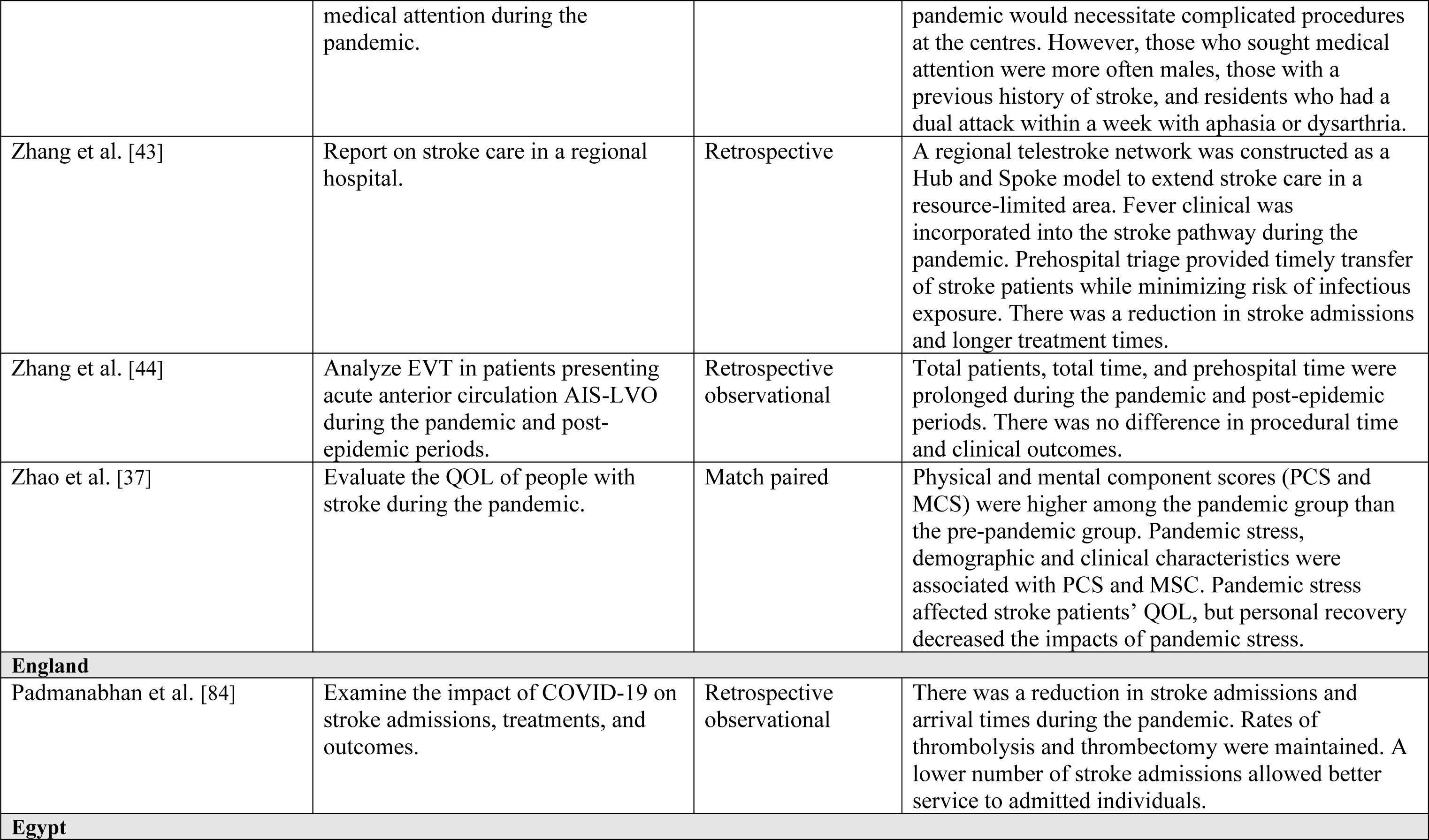
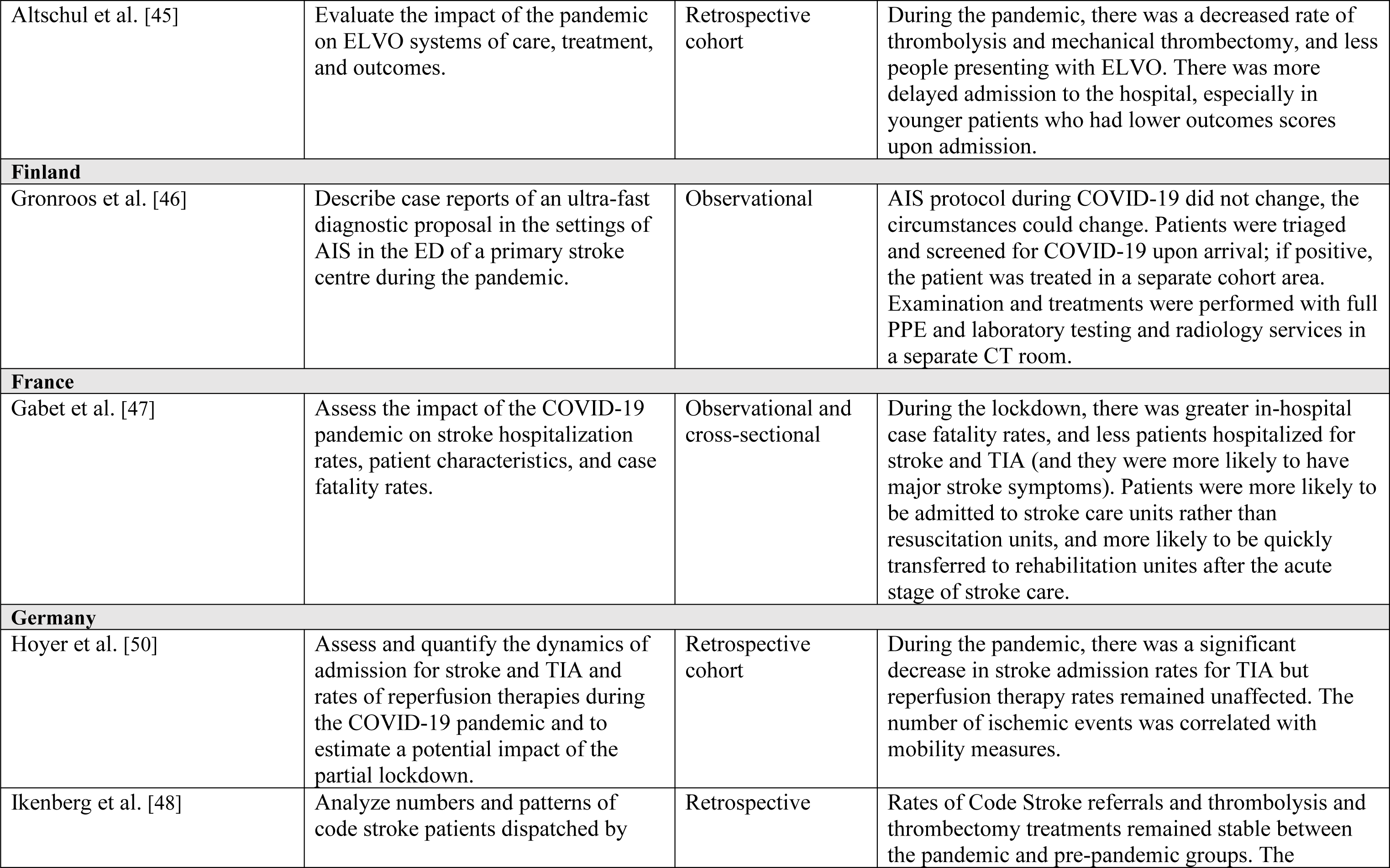
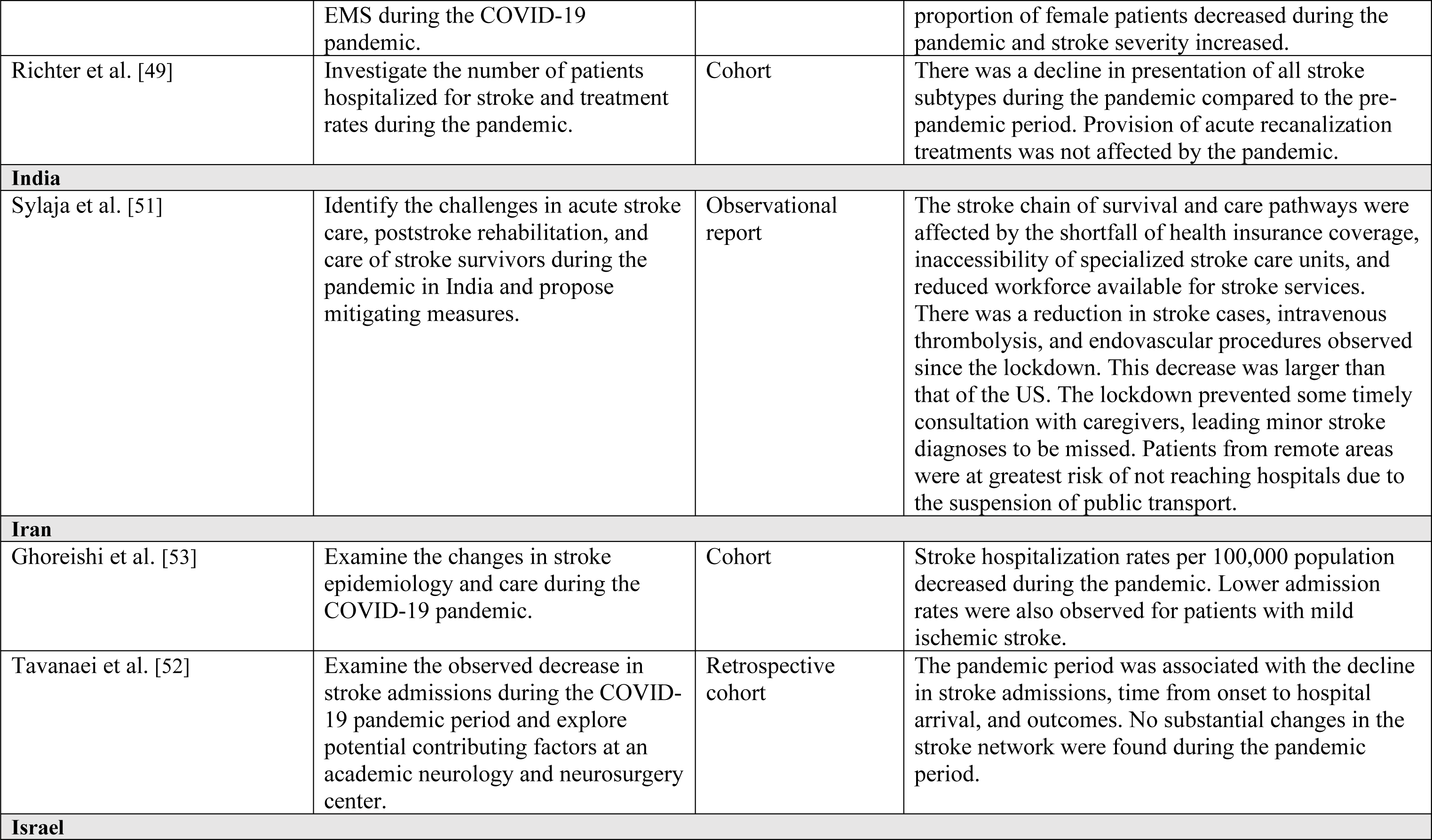
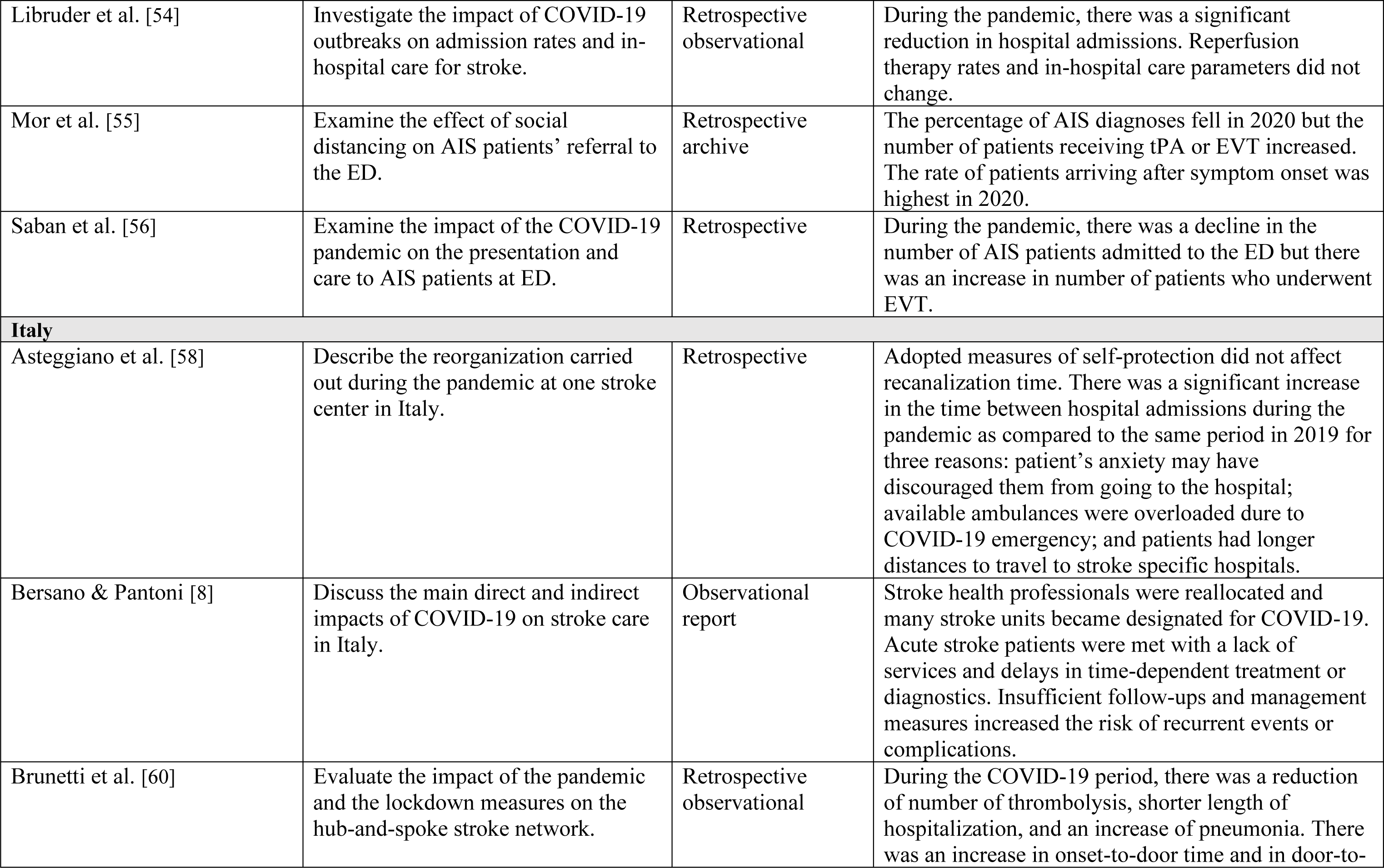
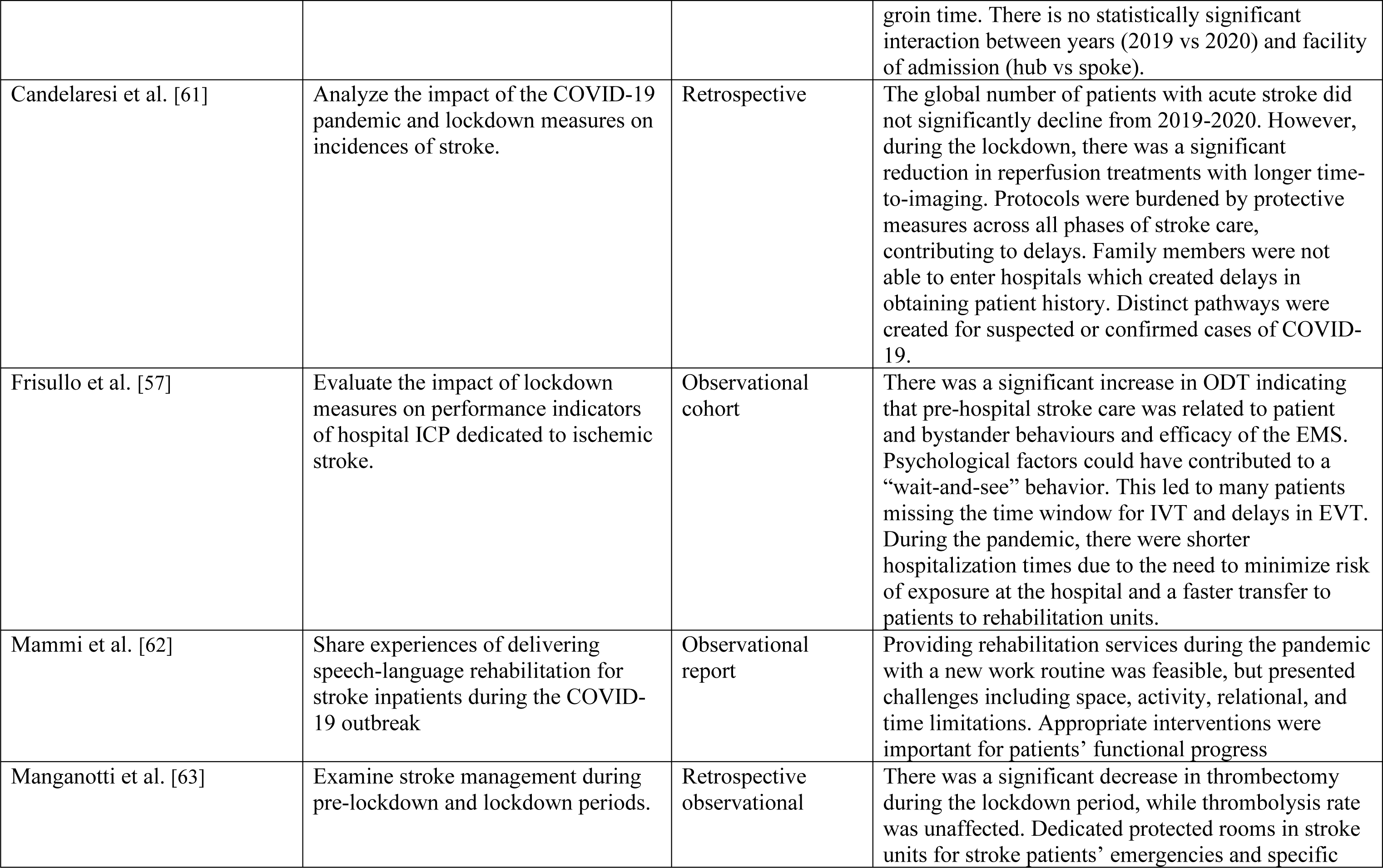
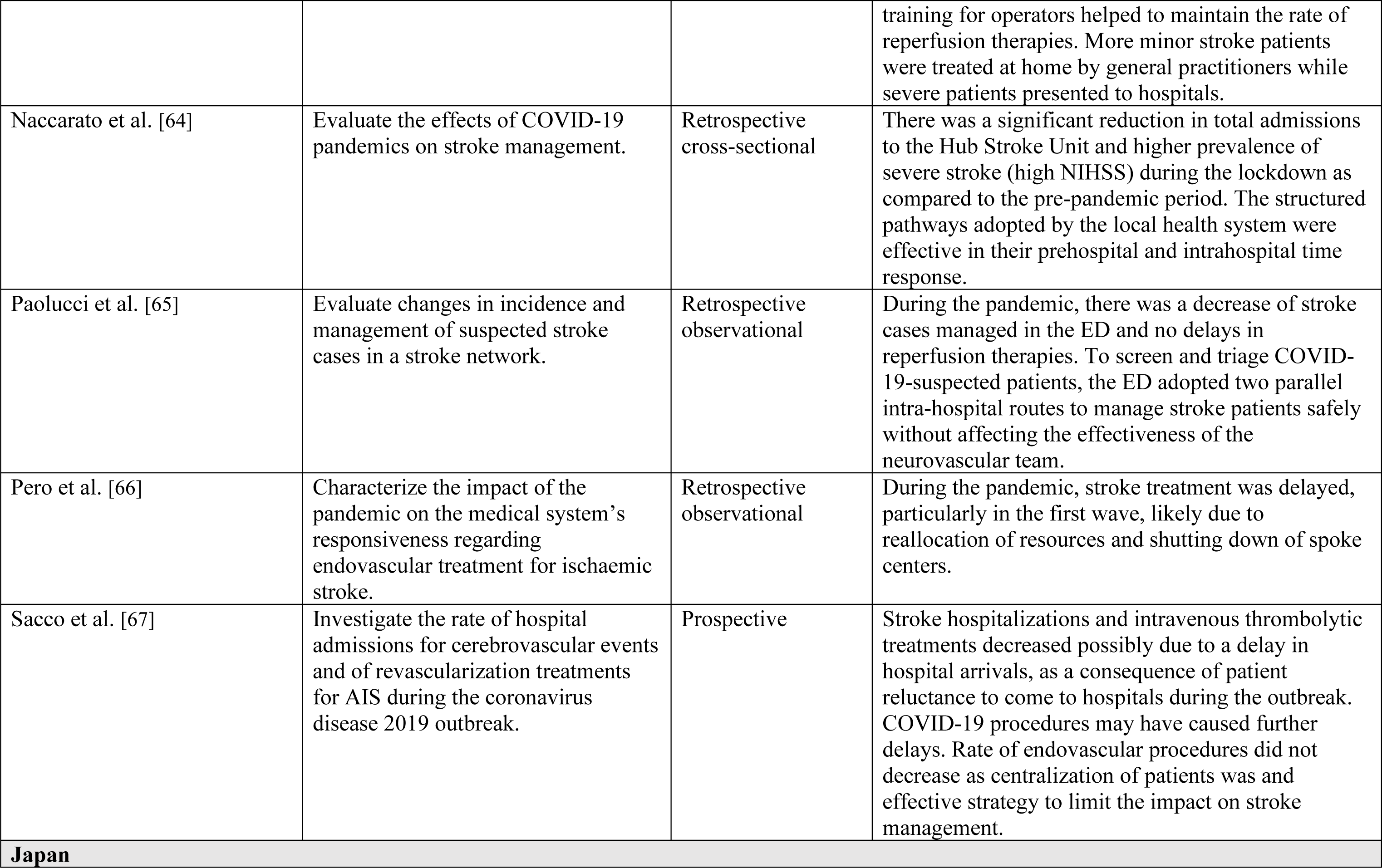
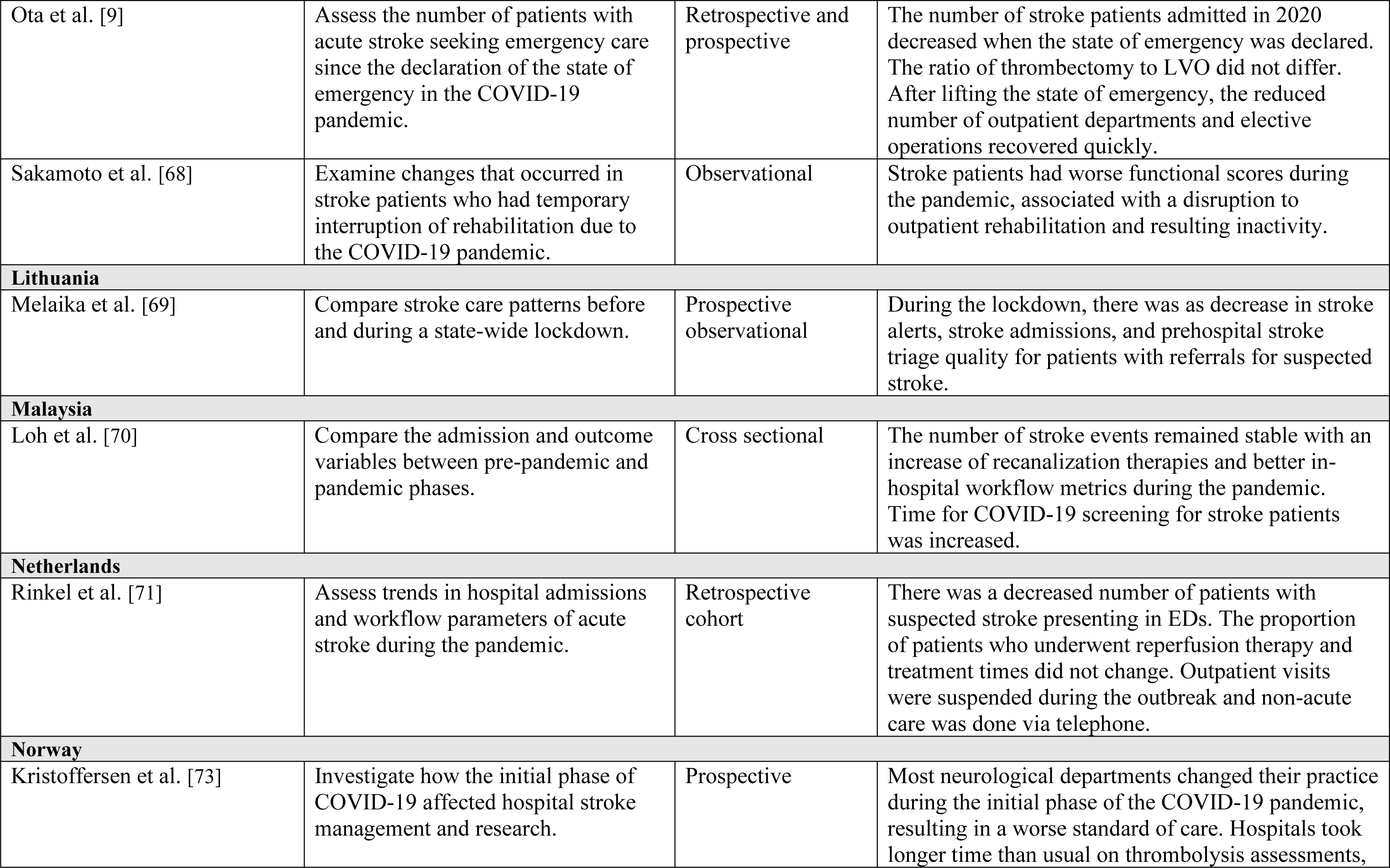
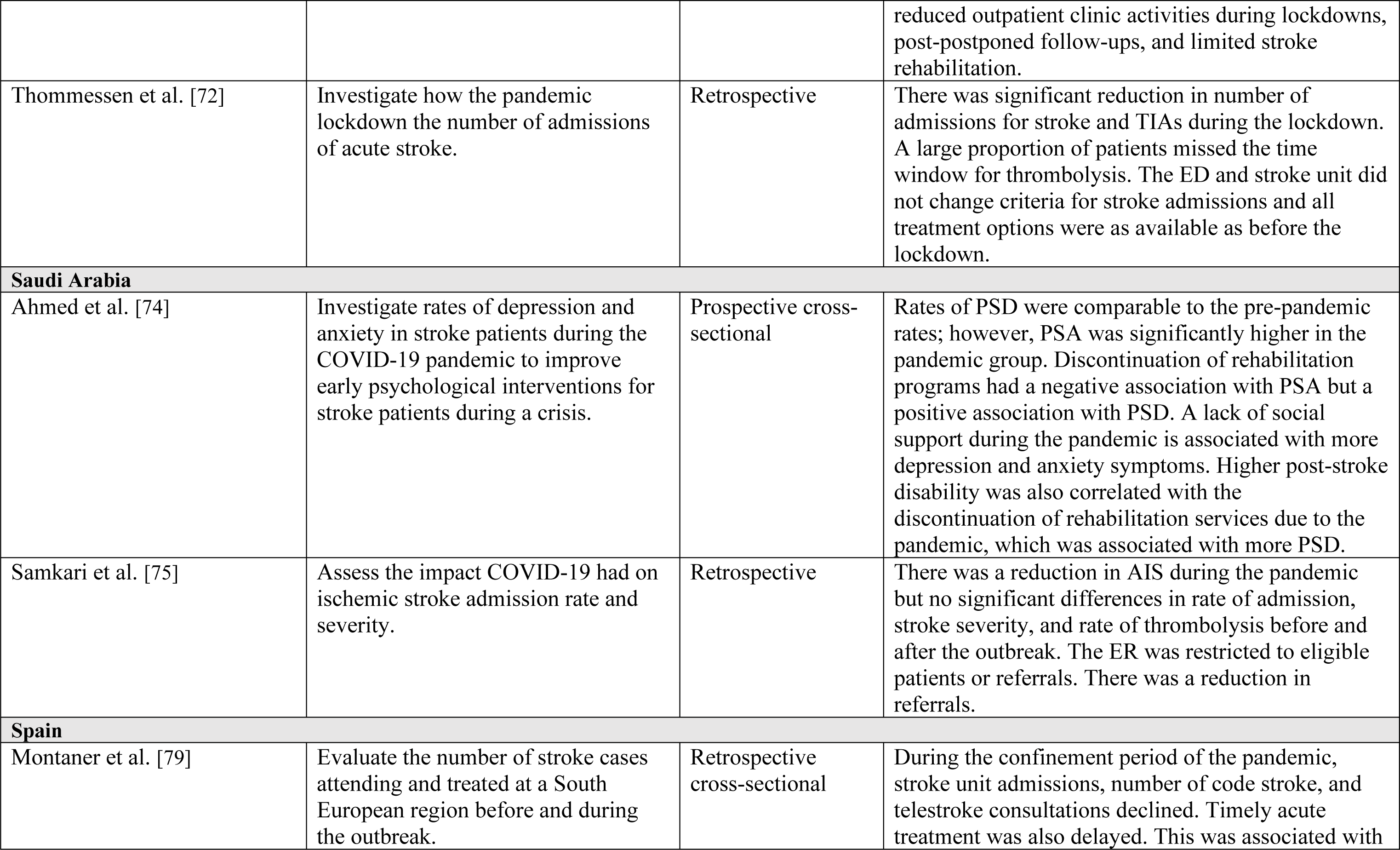
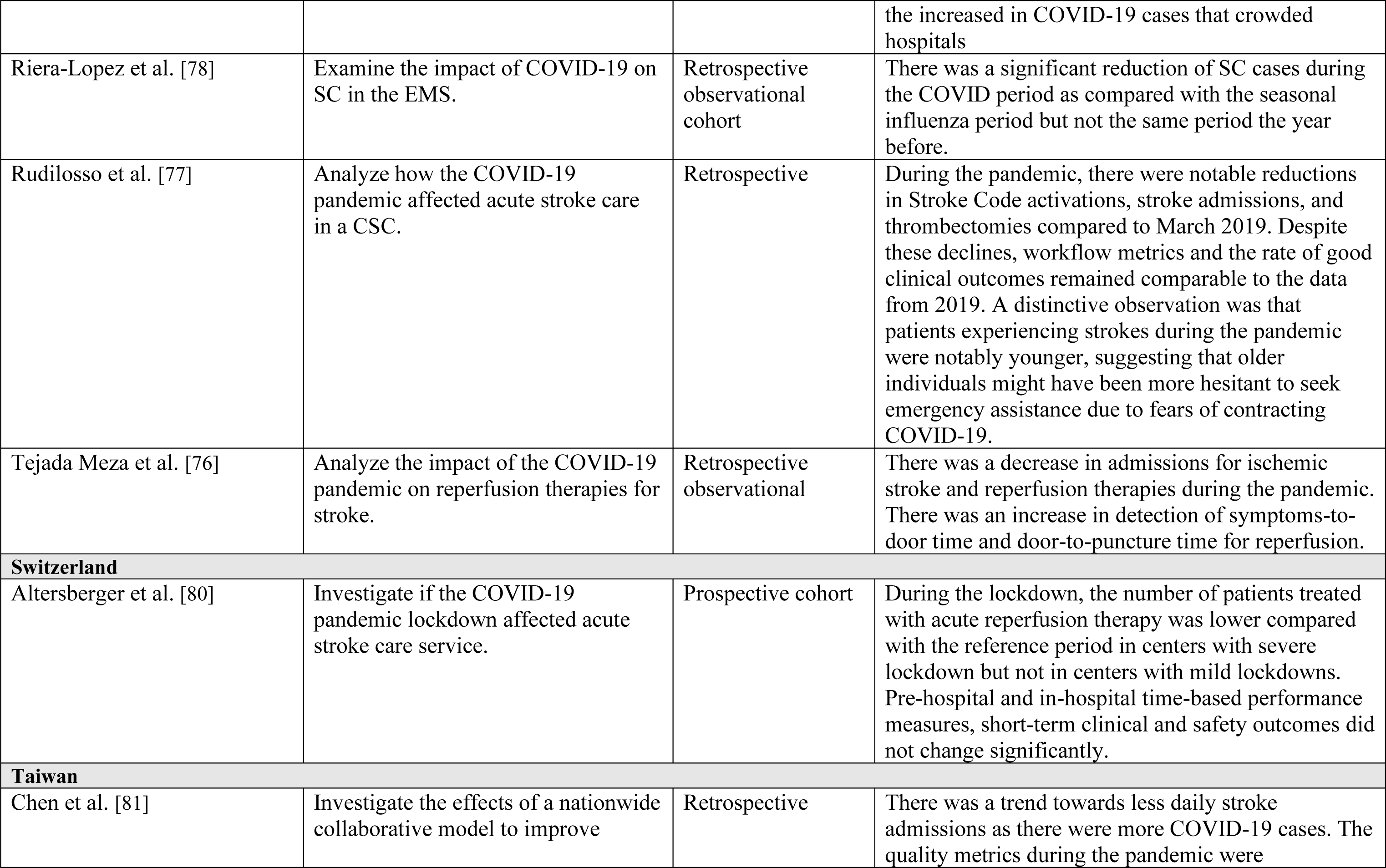
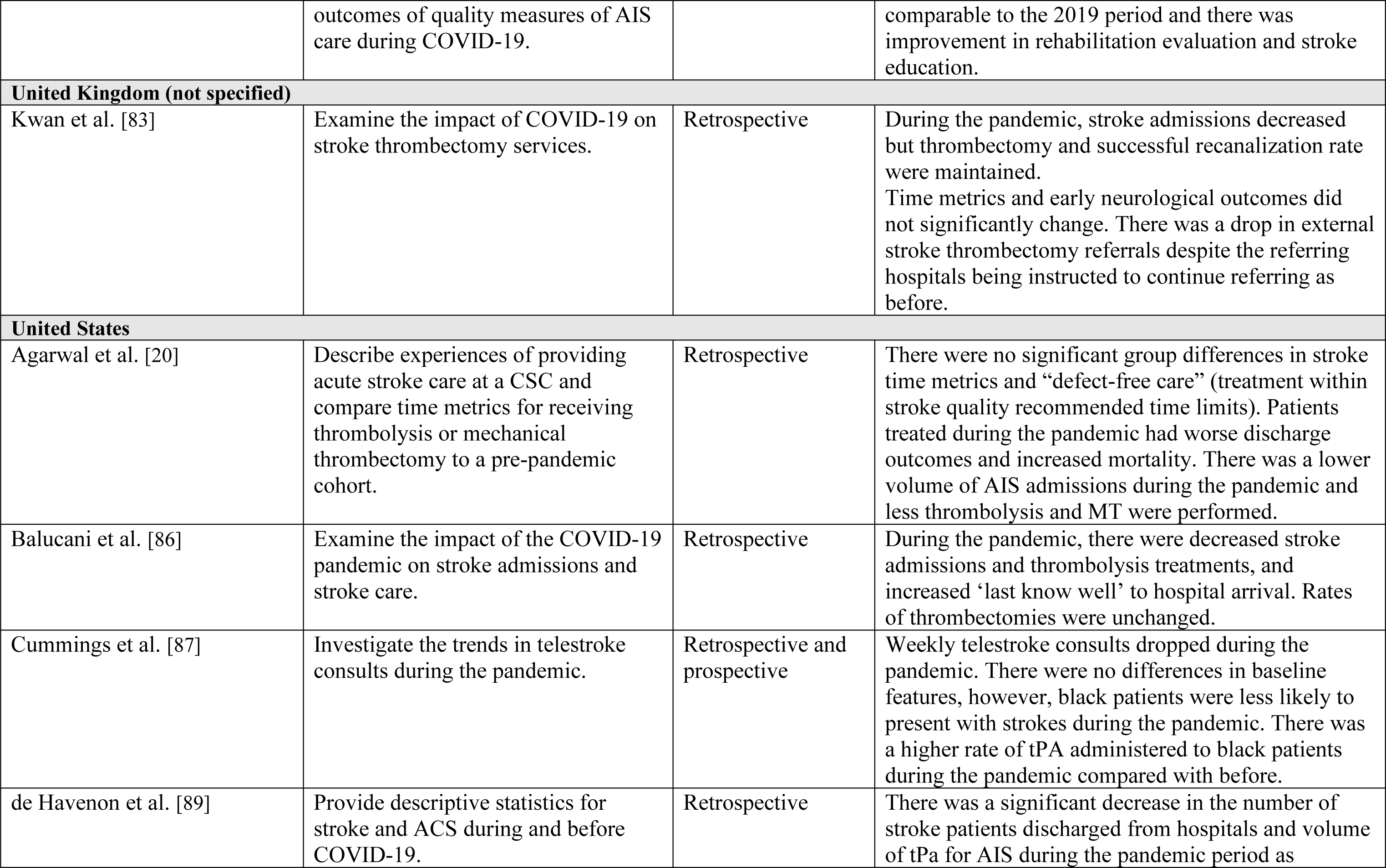
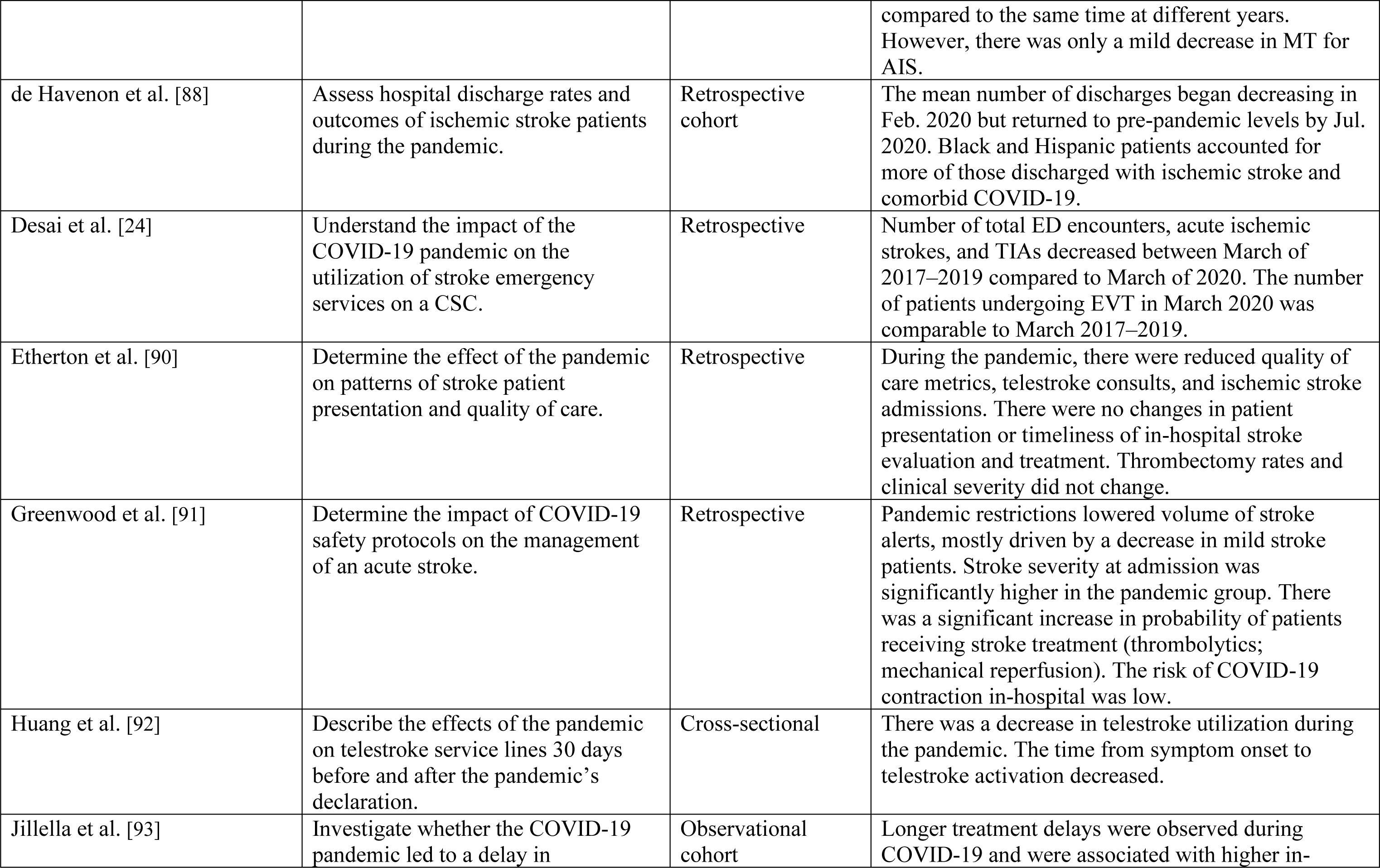
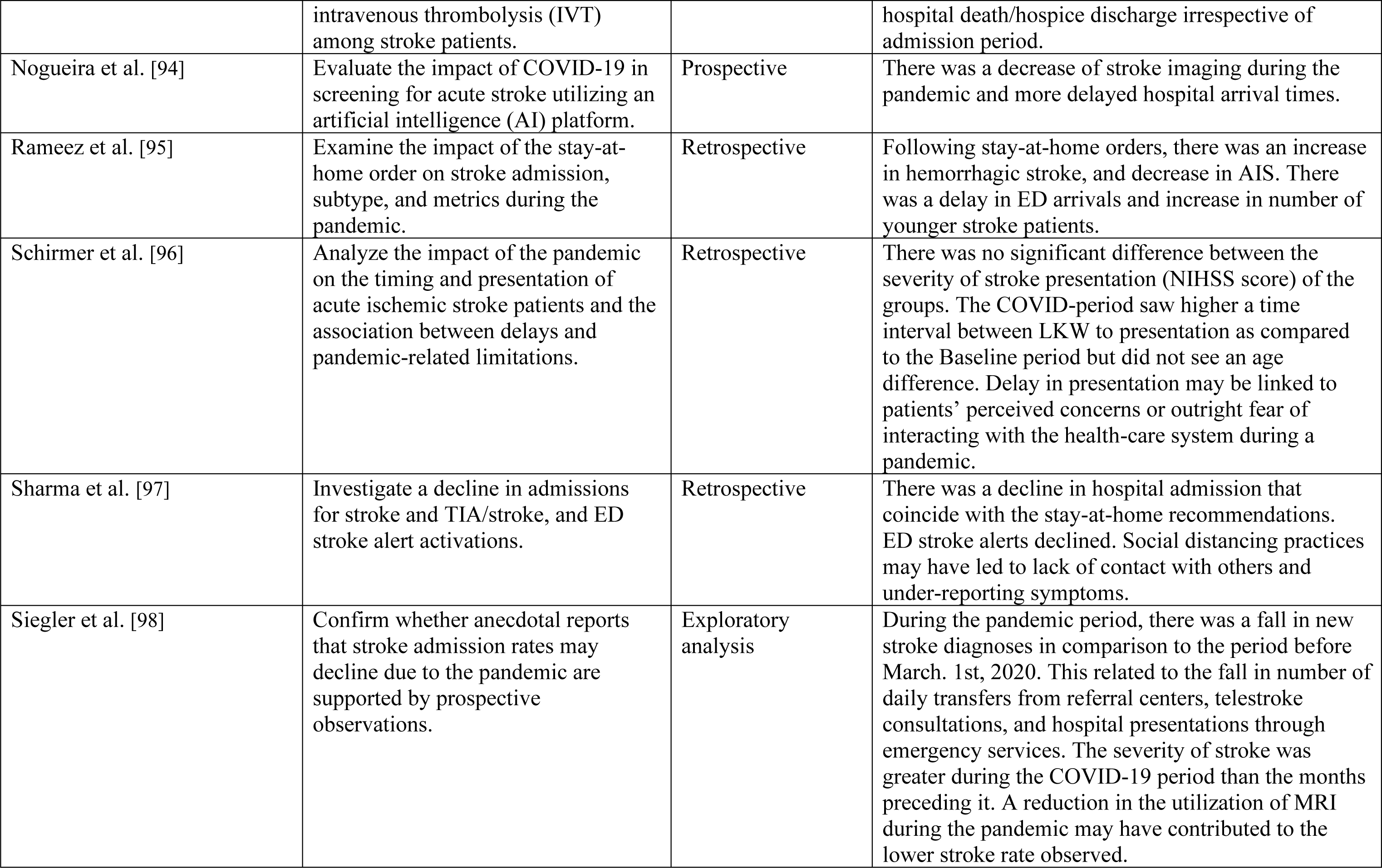
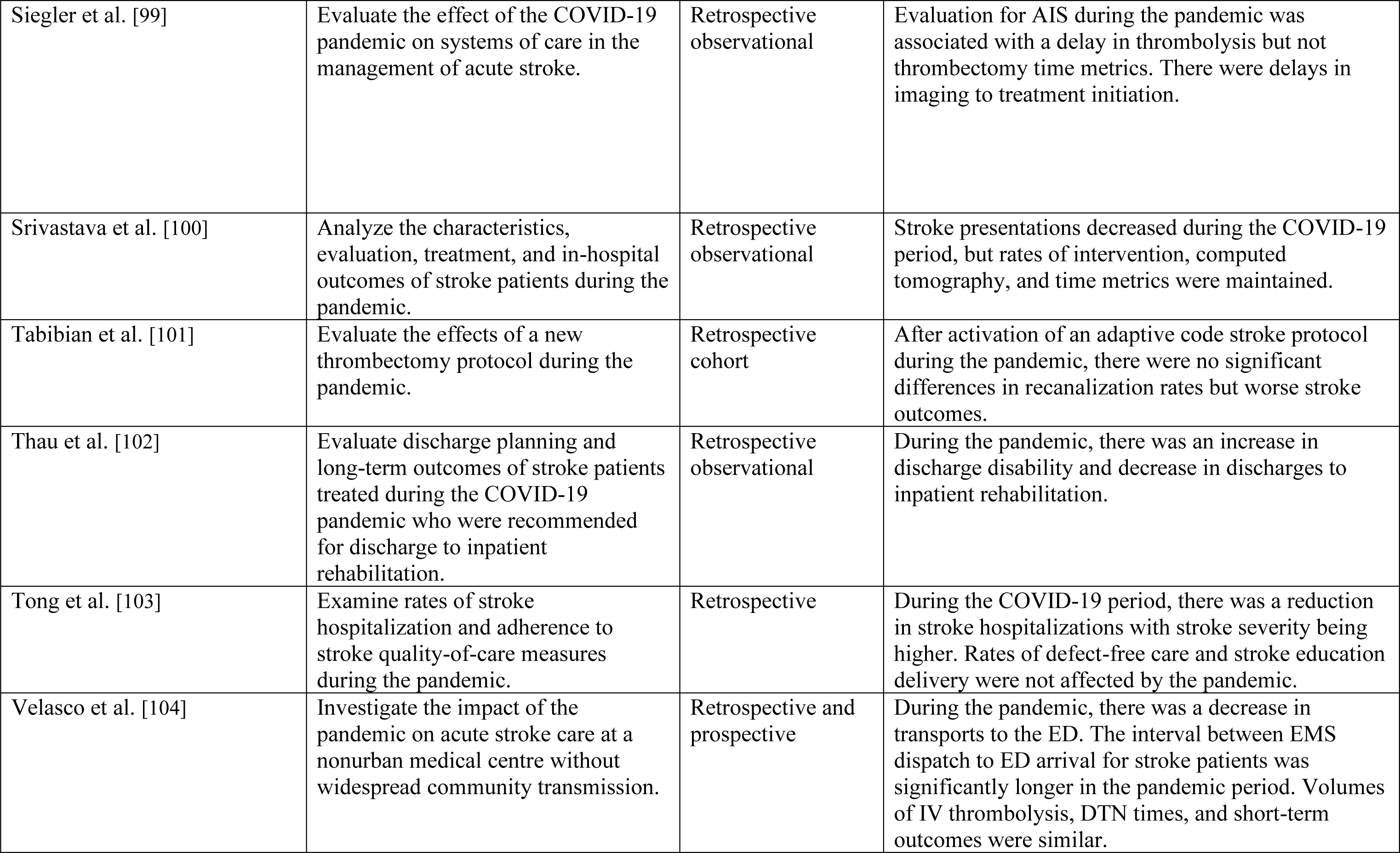
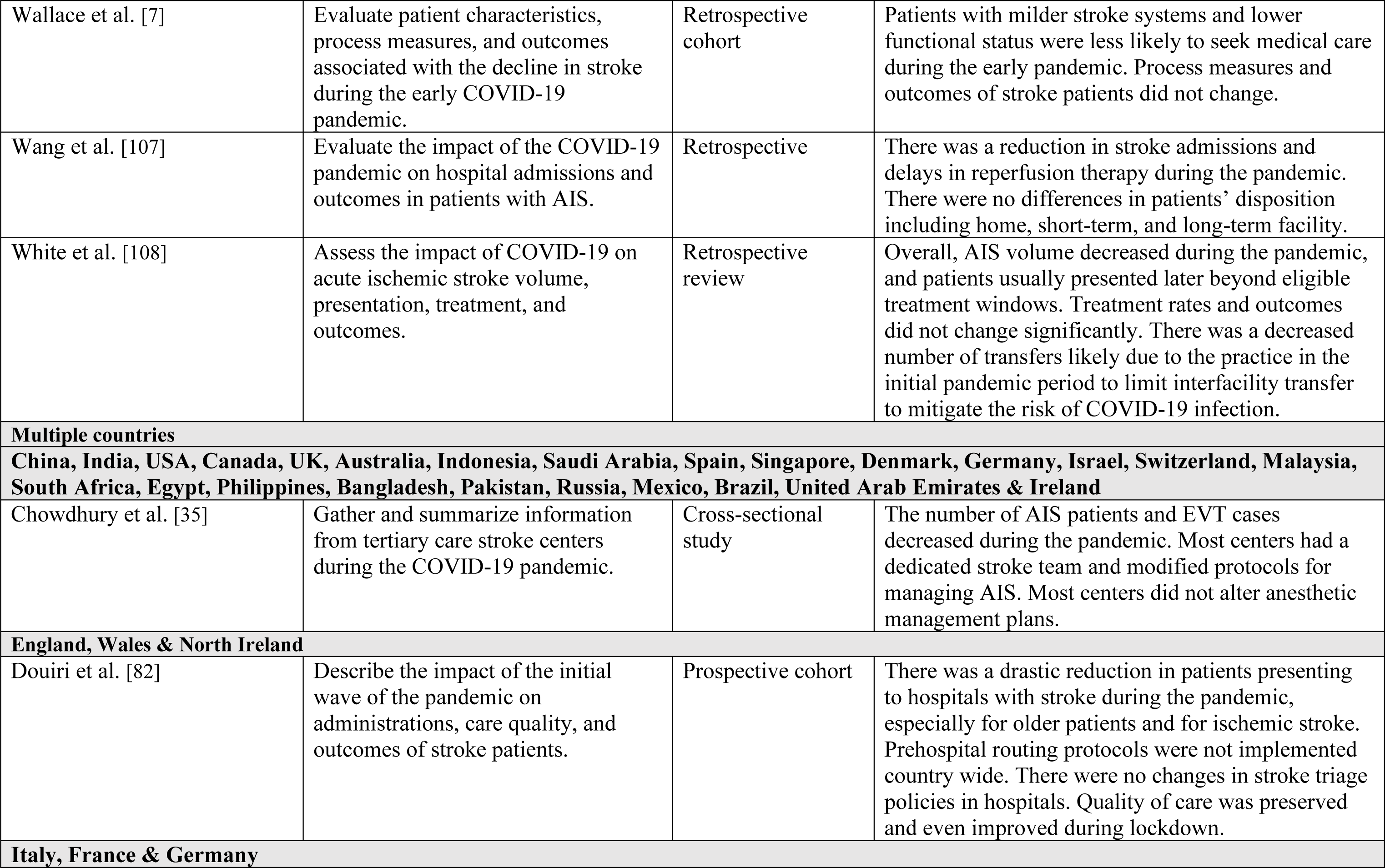
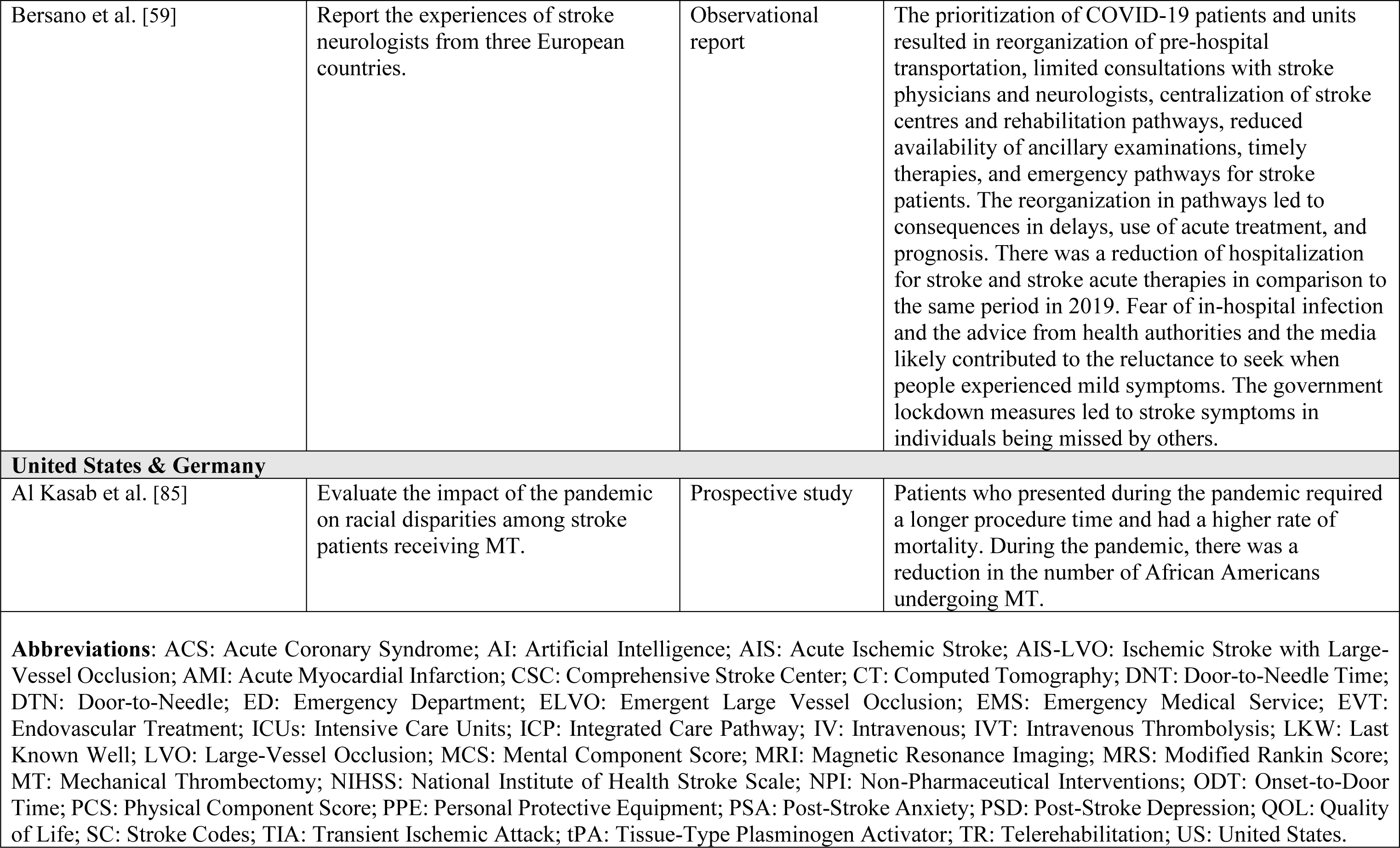

## Non-standard Abbreviations and Acronyms

CT: Computerized Tomography
ELVO: Emergent Large Vessel Occlusion
HCP: Health Care Professional
MERS: Middle East Respiratory Syndrome
NHS: National Health Service
PRISMA-ScR: Preferred Reporting Items for Systematic Reviews and Meta-Analyses extension for Scoping Reviews
TIA: Transient Ischemic Attack

